# Associations between antipsychotic use, substance use and relapse risks in patients with schizophrenia - real-world evidence from two national cohorts

**DOI:** 10.1101/2022.02.09.22270707

**Authors:** Markku Lähteenvuo, Jurjen J. Luykx, Heidi Taipale, Ellenor Mittendorfer-Rutz, Antti Tanskanen, Albert Batalla, Jari Tiihonen

**Affiliations:** Department of Forensic Psychiatry, University of Eastern Finland, Niuvanniemi Hospital, Niuvankuja 65, 70240, Kuopio, Finland; Department of Psychiatry, UMC Utrecht Brain Center, University Medical Center Utrecht, Utrecht University, Utrecht, The Netherlands; Department of Psychiatry and Neuropsychology, School for Mental Health and Neuroscience, Maastricht University Medical Centre, Maastricht, The Netherlands; Second opinion outpatient clinic, GGNet Mental Health, Warnsveld, The Netherlands; Karolinska Institutet, Department of Clinical Neuroscience, Division of Insurance Medicine, Berzelius väg 3. 171 77 Stockholm, Sweden; University of Eastern Finland, School of Pharmacy, Yliopistonranta 1, 70210, Kuopio, Finland; Center for Psychiatry Research, Norra Stationsgatan 69, 11364 Stockholm, Sweden

**Keywords:** antipsychotic, schizophrenia, substance use, clozapine, real-world

## Abstract

**Background:** Research on the effectiveness of pharmacotherapies for schizophrenia-substance use disorder comorbidity (Sch-SUD) is very sparse and non-existent on the prevention of the development of substance use disorders (SUD) in patients with schizophrenia.

**Aims:** To compare the real-world effectiveness of antipsychotics in schizophrenia in decreasing risks of: 1) development of an initial SUD; and 2) psychiatric hospitalization and hospitalization due to SUD among patients with SUD.

**Methods:** Two independent national cohorts including all persons diagnosed with schizophrenia (N=45,476) were followed up for 22 (Finland: 1996–2017) and 11 years (Sweden: 2006–2016). Risks of developing SUD were calculated with between-individual models, and risks of psychiatric and SUD hospitalization with within-individual models using Cox regression and adjusted Hazard Ratios (aHRs) when using vs. not using certain antipsychotics.

**Results:** For schizophrenia patients without SUDs, clozapine use (aHR 0.20, 95%CI 0.16–0.24, p<0.001, in Finland; 0.35, 0.24–0.50, p<0.001, in Sweden) was associated with lowest risks of developing an initial SUD in both countries. Antipsychotic polytherapy was associated with second lowest risk (0.54, 0.44-0.66) in Sweden, and third lowest risk (0.47, 0.42-0.53) in Finland. Risks of psychiatric hospitalization and hospitalization due to SUD were lowest during use of clozapine, antipsychotic polytherapy, and long-acting injectables in both countries. Results were consistent across both countries.

**Conclusion:** Clozapine and antipsychotic polytherapy are most strongly associated with reduced chances of developing SUDs among patients with schizophrenia and with lower relapse rates among patients with both diagnoses.

## Introduction

Schizophrenia is a common^1^ mental disorder that accounts for a tremendous health care burden,^2^ and is frequently comorbid with substance use disorders (SUD)^3^. Substance use not only increases the risk of developing psychotic symptoms, but also negatively affects the course of illness^4,5^. Possible consequences of comorbid substance use include worsening of psychotic symptoms, treatment nonadherence, pharmacodynamic interactions with prescribed agents, increased risk of medical comorbidities, increased violence (both as offenders and victims) and suicides^5–7^. Patients with schizophrenia and comorbid SUD also show increased service utilization (e.g. emergency room visits, hospitalizations) and premature mortality^5–7^. While schizophrenia and SUD are associated with premature death, their comorbidity shows additive effects on mortality. Generally, patients with co-occurring schizophrenia and SUD are considered relatively challenging to treat due to severe positive symptoms, more frequent relapses and frequently observed medication non-compliance^8^.

To our knowledge, the European First Episode Schizophrenia Trial (EUFEST) and Clinical Intervention Trial of Antipsychotics Effectiveness (CATIE) are the largest clinical trials that have investigated SUD subgroup differences in treatment response for patients with schizophrenia. In EUFEST, SUDs were not associated with outcome^9^, while in CATIE moderate substance users showed relatively poor response to antipsychotics^10^. Similarly inconsistent results have been found in several small-scale studies, including small randomized clinical trials, case-reports and small observational studies^11–14^. Although researchers and clinicians have suggested that atypical antipsychotics and, in some cases, clozapine should be preferred for patients with comorbid schizophrenia and SUD (Sch-SUD)^15^, to date there is little evidence to inform prescribing guidelines^16,17^. In a recent systematic review, authors cautioned against the use of conventional LAIs in this patient group, due to risk of increased drug craving, although the authors note that evidence on the use of LAIs in this patient group is still very scarce^18^. Since then, at least one study with a follow-up of one year has investigated the effectiveness of atypical LAIs in this patient group and found favourable effects on quality of life, general functioning (as evaluated with Clinical Global Impression scales) and substance craving^19^. Sch-SUD patients have also been shown to have more EPS-symptoms than patients without SUD^20^, an observation that would be in favor of clozapine, which may decrease chances and severity of EPS. Finally, a recent meta-analysis noted important limitations to the current evidence for the use of antipsychotics in Sch-SUD, highlighting the need for large-scale, good quality studies into this topic^15^.

In sum, research on the effectiveness of pharmacotherapies for Sch-SUD is very sparse, and even more importantly, non-existent on the prevention of the development of SUD in patients with schizophrenia. Therefore, we studied which antipsychotics are associated with the lowest risk of the initial onset of SUD in schizophrenia and what are the most effective treatments for Sch-SUD in preventing relapses. To that end, we collected cohorts totalling more than 45,000 patients with schizophrenia from two independent national registries.

## Methods

### Study cohorts

We had access to two nationwide cohorts of persons with schizophrenia from Finnish and Swedish national registries and refer to them as the Finnish and the Swedish cohorts throughout this manuscript. The Finnish cohort included all persons treated due to schizophrenia (the International Classification of Diseases [ICD] codes F20 and F25 [ICD-10]; and 295 [ICD-8 and-9]) in inpatient care during 1972-2014 in Finland, who entered the cohort at age <46 years. The cohort was identified from the Hospital Discharge register (HDR) maintained by the National Institute of Health and Welfare. Data were collected from the HDR (all hospital care periods with diagnoses, 1972-2017), Prescription register (reimbursed prescription drug purchases, 1995-2017) and causes of death register from Statistics Finland (1972-2017). The follow-up started on January 1, 1996 for persons diagnosed before that, and at the first discharge from inpatient care for persons diagnosed during 1996-2014. The follow-up time ended at death or December 31, 2017, whichever occurred first. This Finnish cohort included 30,860 persons with schizophrenia.

The Swedish cohort included all persons with schizophrenia diagnoses (ICD-10 F20, F25) and registered schizophrenia treatment contact between July 1, 2006 until December 31, 2013 in Sweden, who entered the cohort at age <46 years. Schizophrenia diagnoses were derived from the National Patient Register (NPR, maintained by the National Board of Health and Welfare, inpatient and specialized outpatient care), and the MiDAS register (disability pensions and work disability, maintained by the Swedish Social Insurance Agency). Data were collected from NPR (all hospital care periods and specialized outpatient visits with diagnoses, July 2005-December 2016), the Prescribed Drug Register (PDR, maintained by the National Board of Health and Welfare, prescription drug purchases July 2005-December 2016), the Causes of Death Register (maintained by the National Board of Health and Welfare, causes of death 2006-2016) and the LISA register (maintained by Statistics Sweden, demographic characteristics). The follow-up started on July 1, 2006 for persons diagnosed before that, and at the first recorded diagnoses for persons diagnosed during July 2006-December 2013. The follow-up time ended at death or December 31, 2016, whichever occurred first. The Swedish cohort included 14,616 persons. The cohort and methods have been described previously.^21^

The main differences between the cohorts are the shorter follow-up time for the Swedish cohort (11 years, i.e. years 2006-2016, vs. 22 years, 1996-2017, in the Finnish cohort) and the Finnish cohort only including individuals with a history of a hospitalization due to schizophrenia (the Swedish cohort also included patients identified using diagnoses obtained from disability pensions, sickness absences and specialized outpatient care contacts). The age limit for inclusion was set at <46 years to reduce both chances of survival bias and the number of iatrogenic SUDs arising from benzodiazepine and opioid prescriptions for example to patients undergoing major surgery or starting to suffer from geriatric sleep disorders.

The Regional Ethics Board of Stockholm approved this research project (decision 2007/762–31). Permissions were also granted by pertinent institutional authorities at the Finnish National Institute for Health and Welfare (permission THL/847/5.05.00/2015), the Social Insurance Institution of Finland (65/522/2015), and Statistics Finland (TK53-1042-15). The study was registry-based and no contact was made with the participants of the study; therefore, according to legislation in both countries and as per our previous publications, obtaining informed consent from participants was not required.

#### Substance abuse

SUDs were defined utilizing both inpatient and specialized outpatient care registers as diagnoses F10-F19 excluding F17 (nicotine abuse). SUD was defined categorically: a person was considered as not having a SUD until the first recorded diagnoses, and having SUD ever after the first recording. Some persons had SUD already at cohort entry whereas some persons were transferred from “non-SUD” to SUD group during the follow-up. In the Finnish cohort, the non-SUD group included 22,750 persons and the SUD-group 8,642 persons (2853 new onsets of SUD) during the follow-up. In the Swedish cohort, there were 10,102 persons in the non-SUD group and 4,836 persons (1043 new onsets of SUD) had SUD during the follow-up. The group compositions are shown in the supplement (SFigure 1).

#### Exposure

Antipsychotics were defined as Anatomical Therapeutic Chemical (ATC) classification (ATC) codes N05A (lithium N05AN01 excluded) and further categorized into oral versus LAIs according to their drug formulation (oral antipsychotics referred to in text if not explicitly stated as “LAI”). Polytherapy refers to the concomitant use of two or more antipsychotics. Antipsychotics were categorized as aripiprazole, clozapine, olanzapine, quetiapine, risperidone, other oral, any long-acting injection (LAI) or antipsychotic polytherapy. Drug purchases recorded in the register data were modelled into drug use periods using the PRE2DUP method^22^. The method estimates drug use on a day-by-day basis and is based on the calculation of sliding averages of the daily dose in defined daily dosages (DDD) according to individual drug use patterns and it takes time periods of hospital care into account (when drugs are provided by the caring unit and not recorded in the registers). All exposures were defined time-dependently, i.e. they are updated every time anything changes. Time-dependent or time-varying exposure means that changes in medication use vs. non-use and changes in the medication regimen were followed up and updated in the models. For each time interval, medication treatments were categorized as currently ongoing or not.

#### Outcomes

Outcomes were onset of SUD (diagnosed either in inpatient or specialized outpatient care settings) for patients with schizophrenia but no history of SUD (ICD-10 F10-F19, excl. F17); and psychiatric hospitalization (F20-F29) and SUD hospitalization (F10-F19, excl. F17) for patients with Sch-SUD.

#### Overview of statistical analyses

All statistical analyses were conducted independently in the Finnish and Swedish cohorts. Further details on the analysis models are given and pictured in the Supplemental Methods (SFigures 1 and 2). Results are presented as adjusted hazard ratios (aHRs) with 95% confidence intervals (CIs). The p-values for the analyses were corrected for multiple comparisons on a per graph basis using the Benjamini-Hochberg False Discovery Rate correction with a 0.05 threshold for statistical significance. Correlations between the countries for outcome measures for the different antipsychotic exposures used were calculated using Pearson’s correlation (statistical significance threshold: p<0.05).

#### Statistical analyses to compute risks of initial development of an SUD in patients with schizophrenia (between-individuals model)

Traditional multivariate-adjusted Cox regression models were utilized for analyses of the outcome development of initial SUD. This means that individuals undergoing certain exposures (users of clozapine) were compared with individuals not undergoing this exposure (not using clozapine). This analysis may be affected by bias arising from permanent or long-term individual characteristics if the exposure groups differ with regards to these characteristics (e.g., if users of clozapine are more often male than non-users, this may cause gender-based bias). To reduce chances of such bias, the analyses were therefore adjusted for sex, age at cohort entry, number of previous hospitalizations due to psychosis, time since first schizophrenia diagnosis and continuously updated variables for current versus no use of medications (lipid-modifying agents, opioid analgesics, non-opioid analgesics, anticholinergic anti-Parkinson drugs, prior use of LAI), and continuously updated variables for the following diagnoses: cardiovascular disease, diabetes, asthma/chronic obstructive pulmonary disease (COPD), previous cancer, or previous suicide attempt.

#### Statistical analyses to compute risks of hospitalization in patients with schizophrenia already suffering from SUDs (within-individuals model)

Among persons with Sch-SUD, the outcomes psychiatric hospitalization and SUD hospitalization were analyzed as recurrent events, i.e. events that may happen multiple times for the same person and analyses were conducted with within-individual design using stratified Cox regression^21^. This means that individuals are compared against themselves when undergoing different exposures and can contribute to data sets of different exposures, if they switched their medication regimens during follow-up (e.g., using oral olanzapine for the first 10 months, then 5 months using no medication, then 3 months using clozapine followed by two years of polytherapy with clozapine and aripiprazole). The risks derived from these comparisons within individuals are then pooled for each exposure. Since an individual is compared against him/herself, this analysis method eliminates bias from permanent or long-term characteristics. However, only persons having an outcome event can contribute to within-individual analyses, which leads to a lower n than in between-individual comparisons. The analyses were adjusted for time-varying covariates, which were: sequential order of treatments; use of antidepressants; use of benzodiazepines or Z-drugs; use of mood stabilizers and lithium; and time since cohort entry. For analyses on psychiatric hospitalizations, the most common (5 or 10 depending on analysis) specific antipsychotics were included, in addition to polytherapy and the rest were grouped as “other (FG/SG) orals”. Sensitivity analyses were conducted and are outlined in more detail in the Supplemental Methods.

## Results

### Descriptive statistics

In the Finnish cohort, a total of 30,860 persons with schizophrenia were included, of whom 8,110 (26%) had a diagnosis of SUD (mean age 32.9, SD 7.8 years and 71.9% men for the SUD group; and mean age 33.8, SD 7.8 years and 52.5% men for the non-SUD group). The Swedish cohort (N=14,616) followed a similar pattern, although the prevalence of SUD was slightly higher (31%, N=4514, had SUD) and they were slightly older (mean age 34.3, SD 7.6 and 70.4% men for SUD and mean age 35.2, SD 7.4 and 58.2% men for non-SUD) than in the Finnish cohort (Table 1). A flowchart of the study cohorts and groups is shown in Supplemental (S) Figure 1. Persons without SUD had a slightly higher proportion of their outpatient time spent on antipsychotics (80.3% in the Finnish cohort, 78.7% in the Swedish cohort) than persons with SUD (75.7% in the Finnish cohort, 73.3% in the Swedish cohort).

**Table 1.** Baseline characteristics of persons with and without comorbid substance use disorder (SUD) at the start of follow-up for both cohorts of patients with schizophrenia (persons having first diagnosis of SUD during follow-up are part of the SUD cohort).

|  | Finnish cohort |  |  | Swedish cohort |  |  |
| --- | --- | --- | --- | --- | --- | --- |
|  | No SUD<br>N=22750 | SUD<br>N=8110 | p-value | No SUD<br>N=10102 | SUD<br>N=4514 | p-value |
| Male gender, % (n) | 52.5<br>(11936) | 71.9<br>(5830) | <0.0001 | 58.2<br>(5880) | 70.4<br>(3177) | 0.0030 |
| Mean age (SD) | 33.8 (7.8) | 32.9 (7.8) |  | 35.2 (7.4) | 34.3 (7.6) |  |
| Number of previous psychiatric hospitalizations, % (n) |  |  | <0.0001 |  |  | <0.0001 |
| 1 | 28.0<br>(6380) | 26.1<br>(2115) |  | 41.3<br>(4176) | 35.1<br>(1582) |  |
| 2-3 | 32.5<br>(7394) | 30.1<br>(2440) |  | 22.0<br>(2222) | 20.5<br>(926) |  |
| >3 | 39.5<br>(8976) | 43.8<br>(3555) |  | 36.7<br>(3704) | 44.4<br>(2006) |  |
| Time since first SCH diagnoses, years (n) |  |  | <0.0001 |  |  | <0.0001 |
| ≤1 | 42.9<br>(9748) | 50.7<br>(4108) |  | 51.5<br>(5203) | 52.6<br>(2373) |  |
| >1-5 | 12.2<br>(2774) | 15.3<br>(1243) |  | 13.5<br>(1362) | 17.1<br>(771) |  |
| >5 | 45.0<br>(10228) | 34.0<br>(2759) |  | 35.0<br>(3537) | 30.4<br>(1370) |  |
| Previous suicide attempt, % (n) | 8.4 (1915) | 20.8<br>(1688) | <0.0001 | 6.1 (620) | 21.4<br>(967) | <0.0001 |

### Risk of developing an initial SUD

During the follow-up (median 18.2, IQR 10.0-22.0 in the Finnish cohort; 9.9 years, IQR 6.7-10.5 in the Swedish cohort), 2853 (12.5%) patients of the Finnish non-SUD cohort and 1043 (10.3%) of the Swedish non-SUD cohort were diagnosed with their first SUD. Among persons without SUD, use of clozapine was associated with the lowest risk of developing an initial SUD in both countries (Finland: aHR 0.20, 95% CI 0.16-0.24 and Sweden: aHR 0.35, 95% CI 0.24-0.50). In both countries, also use of aripiprazole (Finland: aHR 0.36, 95% CI 0.24-0.55, Sweden: aHR 0.70, 95% CI 0.51-0.95), antipsychotic polytherapy (Finland: aHR 0.47, 95% CI 0.42-0.53, Sweden: aHR 0.54, 95% CI 0.44-0.66), olanzapine (Finland: aHR 0.49, 95% CI 0.42-0.57, Sweden: aHR 0.67 (0.53-0.84) and any LAI (Finland: aHR 0.62, 95% CI 0.51-0.75, Sweden: aHR 0.70, 95% CI 0.52-0.93) were associated with lower risks of developing an initial SUD, whereas risperidone, quetiapine and other oral antipsychotics showed associations with reduced risks only in the Finnish but not in the Swedish cohort (Figure 1). Of all the specific monotherapies, use of quetiapine was associated with the highest SUD risk. The results were highly consistent between the two countries (Pearson’s r = 0.87, p=0.005, Figure 2A). Clozapine use was also associated with the lowest risk of development of an initial SUD when compared head-to-head to the most common antipsychotic, olanzapine, in the Finnish cohort (aHR 0.30, 95% CI 0.20 – 0.44) and in the Swedish cohort (aHR 0.34, 95% CI 0.12-0.92; SFigure 3). In this comparison analysis, clozapine and polytherapy were the only treatments consistently outperforming olanzapine across countries (SFigure 3).

**Figure 1.**
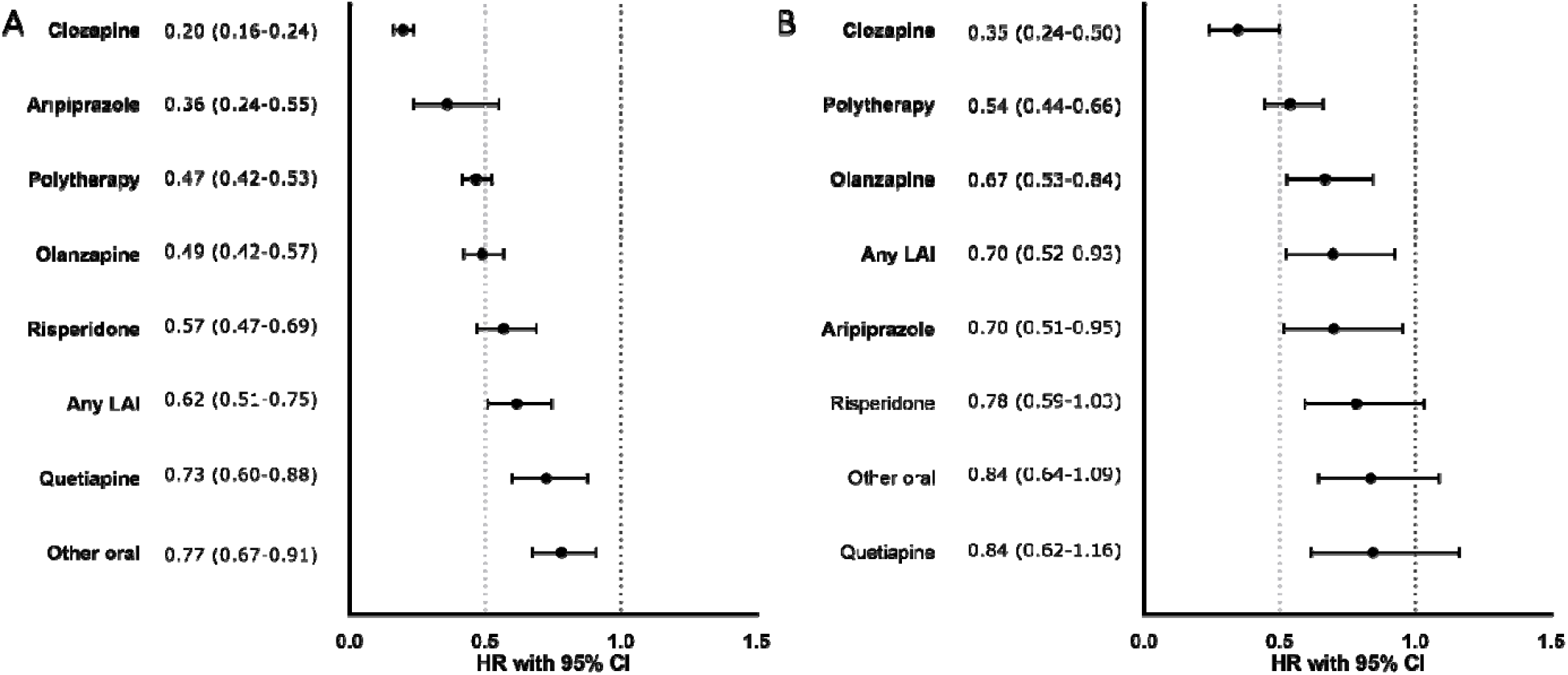
Risks of first SUD (among those without SUD) associated with use of specific antipsychotics, between-individuals model. A) Finnish cohort, B) Swedish cohort. Exposures significant after Benjamini-Hochberg False Discovery Rate correction for multiple comparisons with a 0.05 threshold are bolded. HR= hazard ratio adjusted for covariates.

**Figure 2A:**
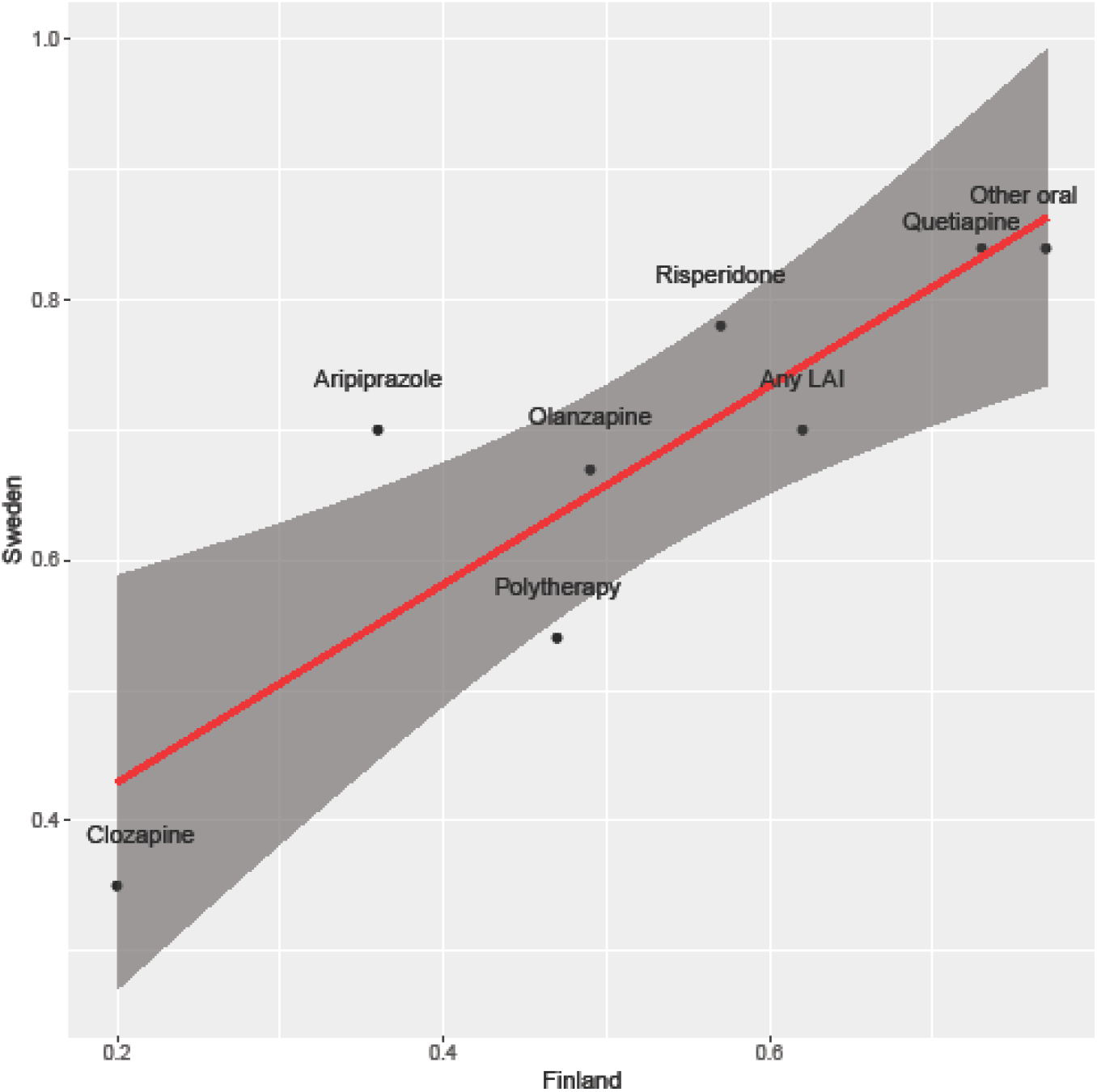
Correlation of effectiveness of antipsychotic treatments (i.e. of adjusted hazard ratios, aHRs) in preventing development of an initial SUD in patients with schizophrenia between the Finnish and Swedish cohorts, Pearson’s r = 0.87, p = 0.005, N=8 antipsychotics. The linear regression curve is depicted with a red line and the 95% confidence interval area for the curve with dark grey. X-axis: aHRs in Finland. Y-axis: aHRs in Sweden.

All sensitivity analyses confirmed the main results (SFigures 4, 5 and 6). The numbers of events and person-years in each treatment category are shown in STable 1.

### Risk of psychiatric hospitalization

Median follow-up time for hospitalizations in persons with SUD was 11.6 years (IQR 6.1-17.7) in the Finnish cohort and 8.6 years (IQR 5.5-10.5) in the Swedish cohort; 5948 persons (73.3%) in the Finnish cohort and 2991 (66.3%) in the Swedish cohort experienced psychiatric hospitalization at least once. In comparison to non-use of antipsychotics (within-individuals model), use of any antipsychotic was associated with a 40% reduction in risk of psychiatric hospitalization in patients with SUD (aHR 0.61, 95% CI 0.58-0.64 for Finland, aHR 0.60, 95% CI 0.56-0.64 for Sweden). For patients with Sch-SUD, risk of psychiatric hospitalization was lowest during use of clozapine (aHR 0.51, 95% CI 0.48-0.54 for Finland, aHR 0.51, 95% CI 0.44-0.58 for Sweden), antipsychotic polytherapy (aHR 0.57, 95% CI 0.54-0.59 for Finland, aHR 0.47, 95% CI 0.44-0.51 for Sweden), any LAI (aHR 0.58, 95% CI 0.54-062 for Finland, aHR 0.67, 95% CI 0.62-0.74 for Sweden) and olanzapine (aHR 0.62, 95% CI 0.58-0.66 for Finland, aHR 0.68, 95% CI 0.61-0.75 for Sweden) compared to non-use (Figure 3, STable1). Use of any specific antipsychotic treatment was associated with reduced risks of psychiatric hospitalization in both countries, but use of quetiapine with the least reduction in risk (aHR 0.83, 95% CI 0.76-0.90 for Finland, aHR 0.81, 95% CI 0.70-094 for Sweden). These beneficial associations of reduced risk for psychiatric hospitalization observed for specific antipsychotic treatments in patients with SUD were similar between the two countries (r=0.84, p=0.009, Figure 2B).

**Figure 2B:**
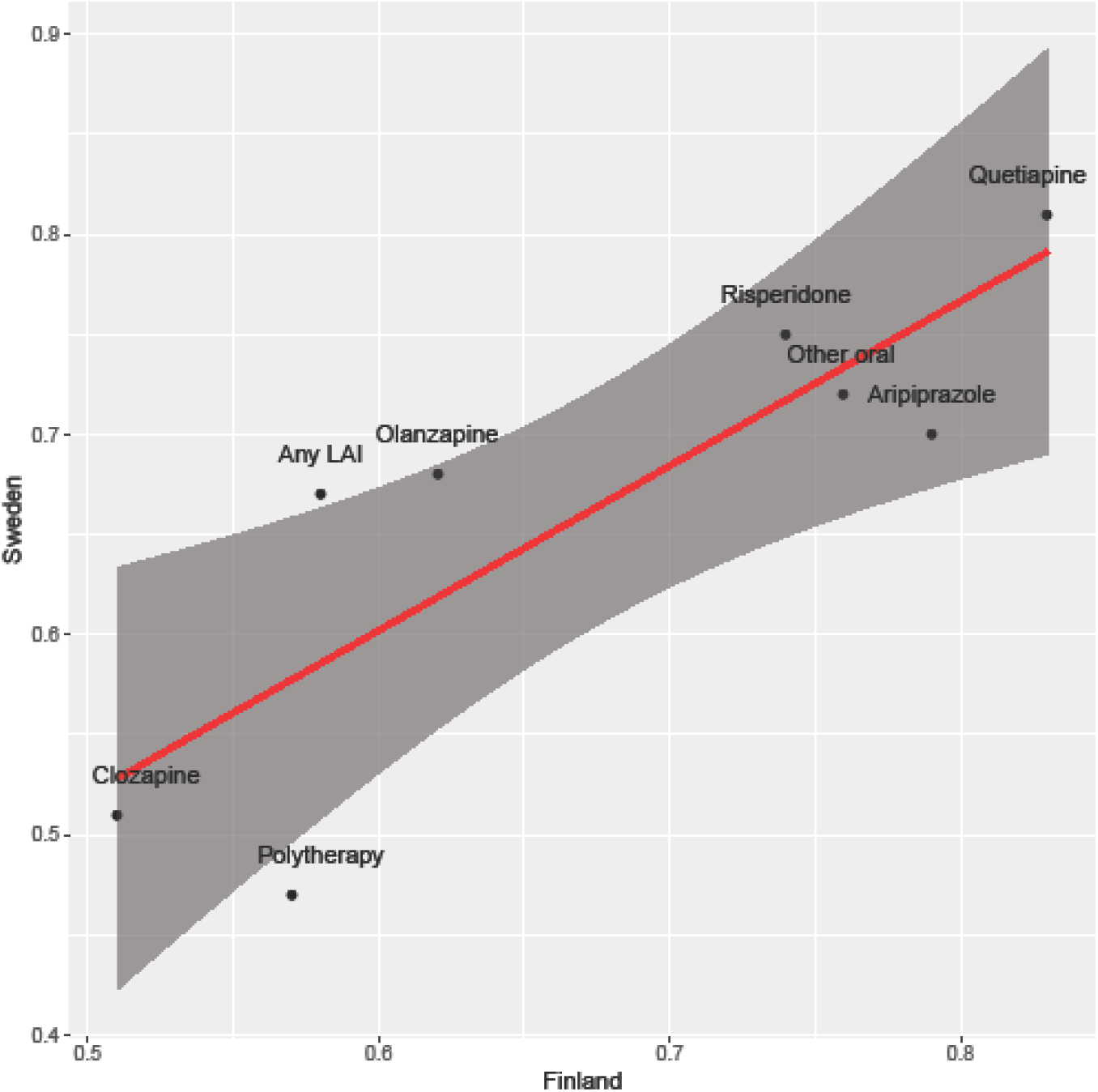
Correlation of effectiveness of antipsychotic treatments (i.e. of adjusted hazard ratios, aHRs) in reducing risk for psychiatric hospitalization in patients with schizophrenia and comorbid SUD between the Finnish and Swedish cohorts, Pearson’s r=0.84, p=0.009, N=8 antipsychotics. The linear regression curve is depicted with a red line and the 95% confidence interval area for the curve with dark grey. X-axis: aHRs in Finland. Y-axis: aHRs in Sweden.

**Figure 3.**
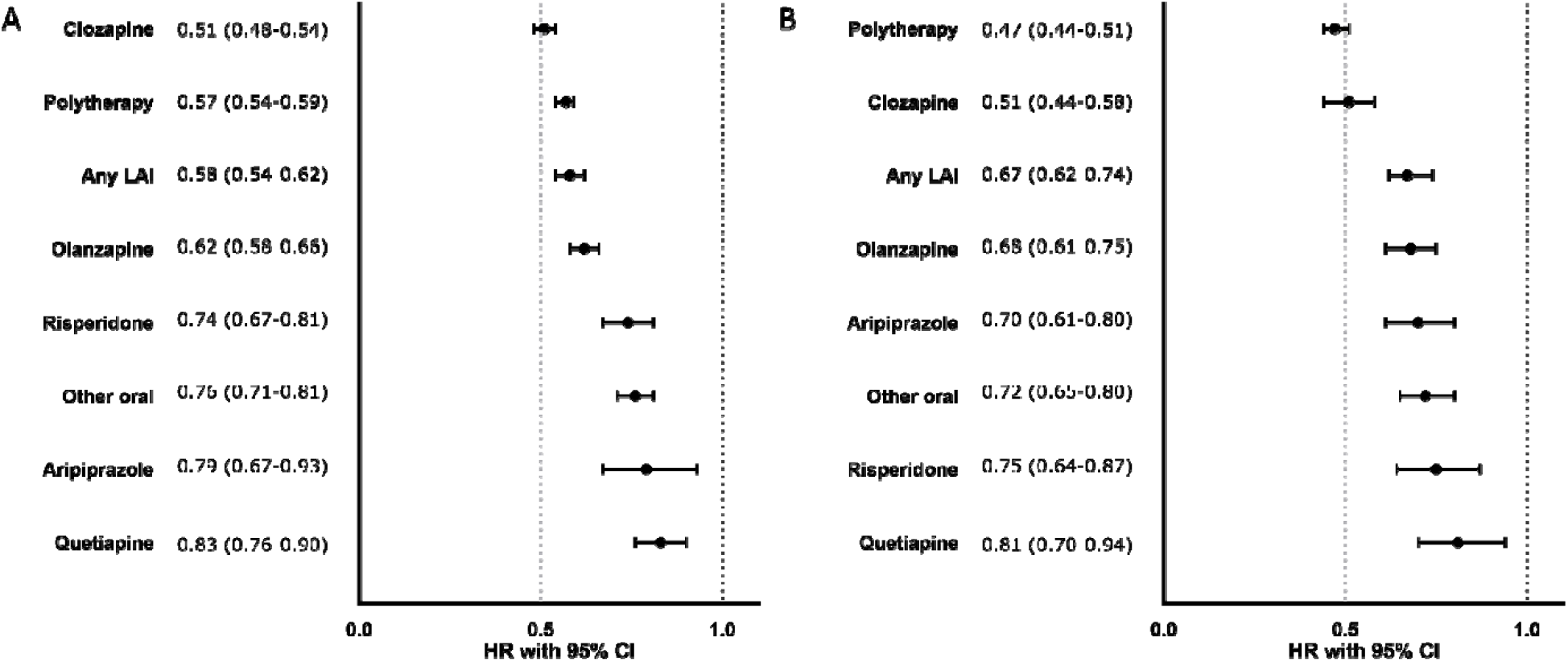
Risks of psychiatric hospitalization associated with use of specific antipsychotics among those with SUD, within-individuals model. A) Finnish cohort, B) Swedish cohort. Exposures significant after Benjamini-Hochberg False Discovery Rate correction for multiple comparisons with a 0.05 threshold are bolded. HR= hazard ratio adjusted for covariates.

### Risk of hospitalization due to SUD

Among persons with SUD, 3971 persons (49.0%) of the Finnish cohort and 2199 persons (48.7%) of the Swedish cohort were hospitalized due to SUD during the follow-up. In individuals with Sch-SUD, use of any antipsychotic was associated with a 19% reduction of SUD hospitalization risk in Finland (aHR 0.81, 95% CI 0.76-0.85) and a 21% reduction in Sweden (aHR 0.79, 95% CI 0.73-0.85) compared with no use of antipsychotics. Of specific treatments, use of clozapine (Finland: aHR 0.59, 95% CI 0.53-0.66; Sweden aHR 0.71, 95% CI 0.54-0.94), antipsychotic polytherapy (Finland: aHR 0.75, 95% CI 0.70-0.79; Sweden: aHR 0.60, 95% CI 0.55-0.66) or LAIs (Finland: aHR 0.79, 95% CI 0.71-0.87; Sweden: aHR 0.84, 95% CI 0.75-0.94) were associated with reduced risks in both countries, whereas olanzapine only in the Finnish cohort (Finland: aHR 0.86, 95% CI 0.78-0.94; Sweden: aHR 0.92, 95% CI 0.82-1.04) (Figure 4 and STable 1). These beneficial associations of reduced risks for hospitalization due to SUD observed for specific antipsychotic treatments in patients with Sch-SUD were similar between the two countries (r=0.71, p=0.049, SFigure 7).

**Figure 4.**
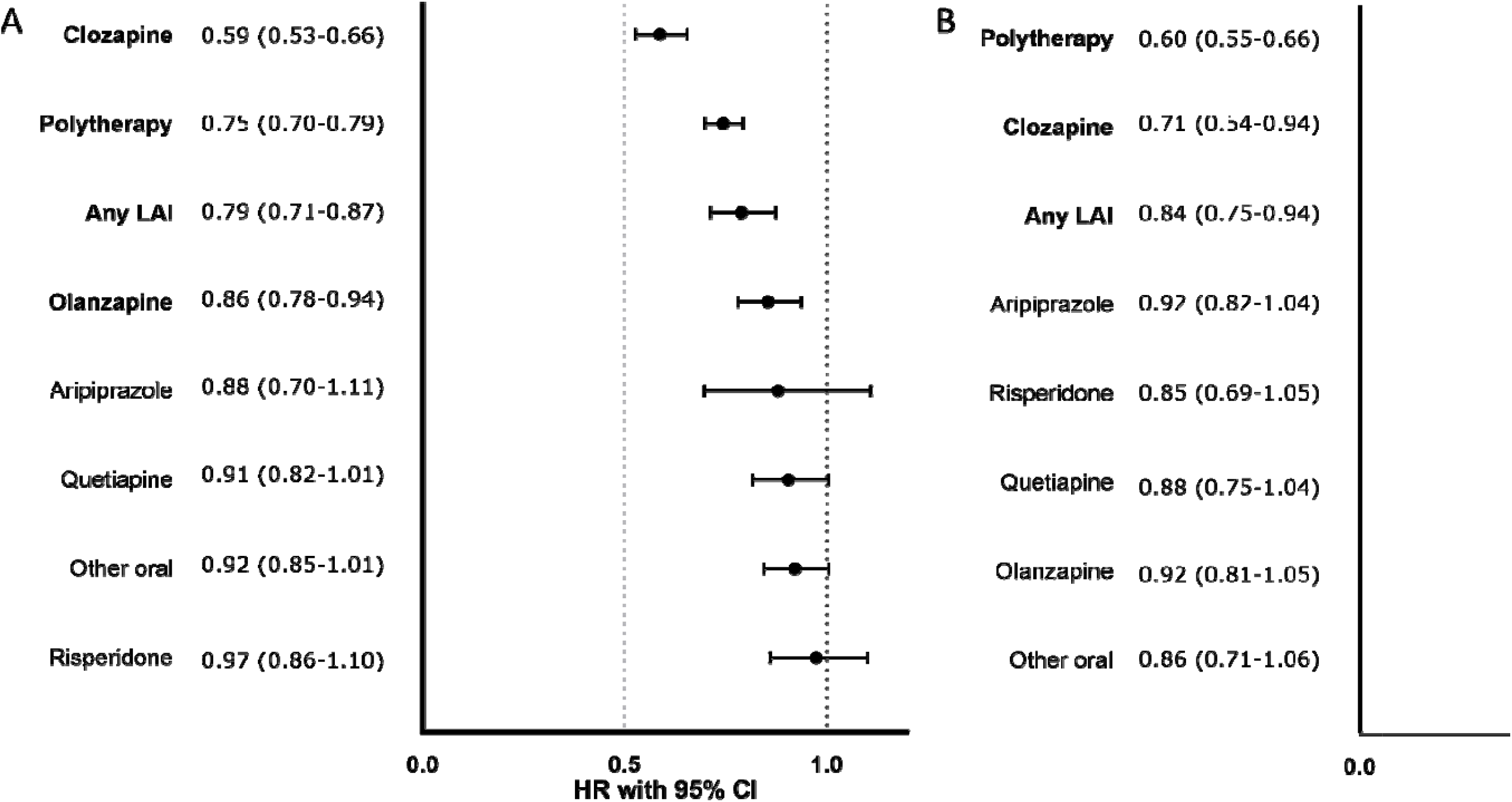
Risks of SUD hospitalization associated with antipsychotic use in those with comorbid substance abuse disorder (SUD), within-individual model, A) Finnish cohort, B) Swedish cohort. HR= hazard ratio adjusted for covariates. Exposures significant after Benjamini-Hochberg False Discovery Rate correction for multiple comparisons with a 0.05 threshold are bolded. HR= hazard ratio adjusted for covariates.

## Discussion

This real-world study provides, for the first time, consistent evidence from two independent cohorts that clozapine use is associated with lower risks of developing SUD in patients with schizophrenia compared to no use or use of other antipsychotics. We also show that in patients with Sch-SUD, clozapine, antipsychotic polytherapy and LAIs are consistently associated with the lowest risks of psychiatric hospitalization and hospitalization due to SUD. Thus, the results give rise to optimism about the treatment of schizophrenia as antipsychotic treatments have now consistently shown to be also effective for those with comorbid SUD.

Our findings are in line with a recent meta-analysis showing superior efficacy of clozapine in Sch-SUD^15^ and other studies pointing towards clozapine’s superiority over other antipsychotics in the treatment of individuals with Sch-SUDs^13^. For example, clozapine has been shown to reduce the subjective effects of cocaine, although it may increase serum cocaine levels, indicating it may be useful in the treatment of cocaine addiction^23^. Others have found that patients with schizophrenia are more likely to remit from alcohol use when using vs. not using clozapine and when using clozapine compared to risperidone^24^.

One possible mechanism explaining our findings relates to the effect some antipsychotics may have on craving. Although not extensively studied, evidence suggests that clozapine reduces craving while other antipsychotic agents have less of an effect on craving^15^. Reduced craving may result in less frequent and lower amounts of substance use in patients with schizophrenia prone to developing SUDs and in patients with Sch-SUD, in turn diminishing their odds of mental and physical symptom worsening and thus of rehospitalization. Alternatively, our findings could be an indication that the self-medication hypothesis of substance use in schizophrenia holds true at least to some extent: possibly, patients on antipsychotics may experience less symptom severity, hence be at lower risk of using drugs of abuse to relieve symptoms and thus of developing SUDs.

Use of clozapine, antipsychotic polytherapy and LAIs were associated with the lowest risks of both SUD and psychiatric hospitalizations among persons with Sch-SUD. The results on polypharmacy are in line with previous results from nationwide cohorts showing a favorable outcome compared with oral monotherapies among persons with schizophrenia in general^21,25^. Possibly, additive effects of antipsychotics in those prescribed antipsychotic polytherapy increase changes of beneficial effects from antipsychotics. Patients with Sch-SUD have been reported to have lower adherence to antipsychotics than other patients with schizophrenia^8^. Among patients with schizophrenia in general, LAIs have been associated with better adherence and lower risk of hospitalizations than their oral counterparts, especially in observational studies^26,27^. However, only a few previous studies examining LAI use among persons with Sch-SUD exist and this literature is inconsistent (see supplement).

### Strengths and limitations

As an observational study, our results are associations and do not prove causality. However, to corroborate our findings we used two individually analyzed nationwide cohorts from two countries with similar, although not identical, healthcare systems and stratified our analyses across different calendar time-periods. The consistent results across main and sensitivity analyses as well as across countries and calendar times lend support to the robustness of our findings and their generalizability to other countries with similar healthcare systems. As the cohorts used were nationwide, the results provided are thus likely to reflect real-world settings. The most significant weaknesses of observational studies are related to confounding by indication. To combat this, the analyses, apart from the analyses of developing SUD where this was impossible, were performed as within-individual analyses, where an individual is used as his/her own control to eliminate bias arising from permanent individual characteristics. Sensitivity analyses were also performed for the between-individual comparisons and the analyses were corrected for a variety of covariates as well as multiple comparisons. However, the registries used do not contain all the information used in clinical decision making, and some residual confounding is bound to remain. For example, we were not able to account for the effects of psychosocial treatment or psychotherapy. Finally, SUDs may sometimes go undiagnosed.

## Conclusions

Overall, our study provides consistent evidence across countries that antipsychotic use in patients with schizophrenia is associated with reduced risks of developing SUDs compared to non-use of antipsychotics. We found that clozapine and antipsychotic polytherapy were most strongly associated with both reduced chances of developing SUDs among patients with schizophrenia and with lower relapse rates in patients with both diagnoses.

## Data Availability

The datasets analysed in this study are not publicly available due to participant privacy and security concerns. Researchers can apply for access to these data from the register holders: for Finnish data with the Social Insurance Institution of Finland (Prescription Register), the Finnish National Institute for Health and Welfare (Hospital Discharge Register), and Statistics Finland (Causes of Death Register); for Swedish data with National Board of Health and Welfare (National Patient Register, Prescribed Drug Register), Statistics Sweden Death and sociodemographic data in LiSA register), and the Swedish Social Insurance Agency (MiDAS register).

## Acknowledgements

We would like to thank ms Aija Räsänen for secretarial assistance.

## Author contributions

All authors conceived the study. HT and AT performed the statistical analyses. ML and JJL wrote the first draft. All authors revised and approved the final draft.

## References

1. Saha S, Chant D, Welham J, McGrath J. A systematic review of the prevalence of schizophrenia. PLoS Medicine. 2005;2(5):0413–0433. doi:10.1371/journal.pmed.0020141

2. Charlson FJ, Ferrari AJ, Santomauro DF, et al. Global epidemiology and burden of schizophrenia: Findings from the global burden of disease study 2016. Schizophrenia Bulletin. 2018;44(6):1195–1203. doi:10.1093/schbul/sby058

3. Hunt GE, Large MM, Cleary M, Lai HMX, Saunders JB. Prevalence of comorbid substance use in schizophrenia spectrum disorders in community and clinical settings, 1990–2017: Systematic review and meta-analysis. Drug and Alcohol Dependence. 2018;191:234–258. doi:10.1016/j.drugalcdep.2018.07.011

4. Heiberg IH, Jacobsen BK, Nesvåg R, et al. Total and cause-specific standardized mortality ratios in patients with schizophrenia and/or substance use disorder. PLoS ONE. 2018;13(8). doi:10.1371/journal.pone.0202028

5. Schmidt LM, Hesse M, Lykke J. The impact of substance use disorders on the course of schizophrenia-A 15-year follow-up study. Dual diagnosis over 15 years. Schizophrenia Research. 2011;130(1-3):228–233. doi:10.1016/j.schres.2011.04.011

6. Dixon L. Dual diagnosis of substance abuse in schizophrenia: Prevalence and impact on outcomes. In: Schizophrenia Research. Vol 35. ; 1999. doi:10.1016/S0920-9964(98)00161-3

7. Margolese HC, Malchy L, Negrete JC, Tempier R, Gill K. Drug and alcohol use among patients with schizophrenia and related psychoses: Levels and consequences. Schizophrenia Research. 2004;67(2-3):157–166. doi:10.1016/S0920-9964(02)00523-6

8. Large M, Mullin K, Gupta P, Harris A, Nielssen O. Systematic meta-analysis of outcomes associated with psychosis and co-morbid substance use. Australian and New Zealand Journal of Psychiatry. 2014;48(5):418–432. doi:10.1177/0004867414525838

9. Wobrock T, Falkai P, Schneider-Axmann T, et al. Comorbid substance abuse in first-episode schizophrenia: Effects on cognition and psychopathology in the EUFEST study. Schizophrenia Research. 2013;147(1):132–139. doi:10.1016/j.schres.2013.03.001

10. Kerfoot KE, Rosenheck RA, Petrakis IL, et al. Substance use and schizophrenia: Adverse correlates in the CATIE study sample. Schizophrenia Research. 2011;132((2-3):177–182. doi:10.1016/j.schres.2011.07.032

11. Buckley P, Thompson P, Way L, Meltzer HY. Substance abuse among patients with treatment-resistant schizophrenia: Characteristics and implications for clozapine therapy. American Journal of Psychiatry. 1994;151(3):385–389. doi:10.1176/ajp.151.3.385

12. Conley RR, Kelly DL, Gale EA. Olanzapine response in treatment-refractory schizophrenic patients with a history of substance abuse. Schizophrenia Research. 1998;33((1-2):95–101. doi:10.1016/S0920-9964(98)00062-0

13. Lee ML, Dickson RA, Campbell M, Oliphant J, Gretton H, Dalby JT. Clozapine and substance abuse in patients with schizophrenia [4]. Canadian Journal of Psychiatry. 1998;43(8):855–856.

14. Zimmet S v., Strous RD, Burgess ES, Kohnstamm S, Green AI. Effects of clozapine on substance use in patients schizophrenia and schizoaffective disorder: A retrospective survey. Journal of Clinical Psychopharmacology. 2000;20(1):94–98. doi:10.1097/00004714-200002000-00016

15. Krause M, Huhn M, Schneider-Thoma J, Bighelli I, Gutsmiedl K, Leucht S. Efficacy, acceptability and tolerability of antipsychotics in patients with schizophrenia and comorbid substance use. A systematic review and meta-analysis. European Neuropsychopharmacology. 2019;29(1):32–45. doi:10.1016/j.euroneuro.2018.11.1105

16. Ziedonis DM, Smelson D, Rosenthal RN, et al. Improving the care of individuals with schizophrenia and substance use disorders: Consensus recommendations. Journal of Psychiatric Practice. 2005;11(5):315–339. doi:10.1097/00131746-200509000-00005

17. Clerici M, de Bartolomeis A, de Filippis S, et al. Patterns of management of patients with dual disorder (Psychosis) in Italy: A survey of psychiatrists and other physicians focusing on clinical practice. Frontiers in Psychiatry. 2018;9. doi:10.3389/fpsyt.2018.00575

18. Zhornitsky S, Stip E. Oral versus Long-Acting Injectable Antipsychotics in the Treatment of Schizophrenia and Special Populations at Risk for Treatment Nonadherence: A Systematic Review. Schizophrenia Research and Treatment. 2012;2012:1–12. doi:10.1155/2012/407171

19. Cuomo I, Kotzalidis GD, de Persis S, et al. Head-to-head comparison of 1-year aripiprazole long-acting injectable (LAI) versus paliperidone LAI in comorbid psychosis and substance use disorder: Impact on clinical status, substance craving, and quality of life. Neuropsychiatric Disease and Treatment. 2018;14:1645–1656. doi:10.2147/NDT.S171002

20. Potvin S, Pampoulova T, Mancini-Marië A, Lipp O, Bouchard RH, Stip E. Increased extrapyramidal symptoms in patients with schizophrenia and a comorbid substance use disorder. Journal of Neurology, Neurosurgery and Psychiatry. 2006;77(6):796–798. doi:10.1136/jnnp.2005.079228

21. Tiihonen J, Mittendorfer-Rutz E, Majak M, et al. Real-world effectiveness of antipsychotic treatments in a nationwide cohort of 29 823 patients with schizophrenia. JAMA Psychiatry. 2017;74(7):686–693. doi:10.1001/jamapsychiatry.2017.1322

22. Tanskanen A, Taipale H, Koponen M, et al. From prescription drug purchases to drug use periods A second generation method (PRE2DUP). BMC Medical Informatics and Decision Making. 2015;15(1). doi:10.1186/s12911-015-0140-z

23. Farren CK, Hameedi FA, Rosen MA, Woods S, Jatlow P, Kosten TR. Significant interaction between clozapine and cocaine in cocaine addicts. Drug and Alcohol Dependence. 2000;59(2):153–163. doi:10.1016/S0376-8716(99)00114-3

24. Green AI, Burgess ES, Dawson R, Zimmet S v., Strous RD. Alcohol and cannabis use in schizophrenia: Effects of clozapine vs. risperidone. Schizophrenia Research. 2003;60(1):81–85. doi:10.1016/S0920-9964(02)00231-1

25. Tiihonen J, Taipale H, Mehtälä J, Vattulainen P, Correll CU, Tanskanen A. Association of Antipsychotic Polypharmacy vs Monotherapy with Psychiatric Rehospitalization among Adults with Schizophrenia. JAMA Psychiatry. 2019;76(5):499–507. doi:10.1001/jamapsychiatry.2018.4320

26. Kishimoto T, Nitta M, Borenstein M, Kane JM, Correll CU. Long-acting injectable versus oral antipsychotics in schizophrenia: A systematic review and meta-analysis of mirror-image studies. Journal of Clinical Psychiatry. 2013;74(10):957–965. doi:10.4088/JCP.13r08440

27. Kane JM, Kishimoto T, Correll CU. Non-adherence to medication in patients with psychotic disorders: Epidemiology, contributing factors and management strategies. World Psychiatry. 2013;12(3):216–226. doi:10.1002/wps.20060

