## supplements for "Associations between antipsychotic use, substance use and relapse risks in patients with schizophrenia - real-world evidence from two national cohorts"

### Supplemental Methods

#### Data-analyses

The within-individual analyses utilized here are described in more detail elsewhere.[1]

#### Sensitivity analyses

As sensitivity analyses, first the risk of initial development of SUD was analyzed by censoring the first 30 days of exposure time from the beginning of each exposure period as this may represent a time period of sub-optimal pharmacological effect, thus correcting for protopathic bias. Second, the risk of first SUD was also analyzed by excluding persons with disorganized/hebephrenic schizophrenia (ICD-10 code F20.1) ever during the follow-up, because those patients may sometimes be too ill to acquire substances of abuse or develop SUD, but are often treated with clozapine, which might have resulted in clozapine seeming more favorable than the other treatment regimens. Third, the analyses on the risk of initial development of SUD was restricted to persons ever using olanzapine (N=7755 in the Finnish cohort and N=3718 in the Swedish cohort) and head-to-head analyses were conducted with olanzapine use as the reference. Head-to-head analyses aim to inform about the risk in reference to most commonly prescribed antipsychotic, allowing between-drug comparisons.

### Supplemental Discussion

A previous study highlights that patients receiving LAIs, both with and without SUD comorbidity, experience less treatment failures and longer time to treatment failure than patients using oral antipsychotics[2]. Furthermore, first-episode patients with comorbid SUDs treated with LAIs experience fewer relapses than those treated with oral counterparts, even though they carried worse prognostic factors, such as history of homelessness[3]. This may be due to patients with schz-SUD having lower adherence towards antipsychotic medications[4,5] and LAIs improving adherence. Thus, the reduced risk of hospitalization during use of LAIs observed in this study is mostly in line with previous literature. However, meta-analyses of randomized clinical trials in patients with schizophrenia have failed to observe a beneficial effect of LAIs over their oral counterparts[6], although more recent studies with first episode patients have shown robust effects in favor of LAIs[7,8]. Also, previous studies have shown clozapine to be superior to other oral agents in patients with sch-SUD[9–11]. This may be due to clozapine’s effect on reducing craving[12,13].

#### References

1. Lichtenstein P, Halldner L, Zetterqvist J, et al. Medication for Attention Deficit–Hyperactivity Disorder and Criminality. *N Engl J Med*. 2012;367(21):2006-2014. doi:10.1056/nejmoa1203241
2. Lynn Starr H, Bermak J, Mao L, Rodriguez S, Alphs L. Comparison of long-acting and oral antipsychotic treatment effects in patients with schizophrenia, comorbid substance abuse, and a history of recent incarceration: An exploratory analysis of the PRIDE study. *Schizophr Res*. 2018;194:39-46. doi:10.1016/j.schres.2017.05.005
3. Abdel-Baki A, Thibault D, Medrano S, et al. Long-acting antipsychotic medication as first-line treatment of first-episode psychosis with comorbid substance use disorder. *Early Interv Psychiatry*. 2020;14(1):69-79. doi:10.1111/eip.12826
4. Lacro JP, Dunn LB, Dolder CR, Leckband SG, Jeste D V. Prevalence of and risk factors for medication nonadherence in patients with schizophrenia: A comprehensive review of recent literature. *J Clin Psychiatry*. 2002;63(10):892-909. doi:10.4088/JCP.v63n1007
5. Kane JM, Kishimoto T, Correll CU. Non-adherence to medication in patients with psychotic disorders: Epidemiology, contributing factors and management strategies. *World Psychiatry*. 2013;12(3):216-226. doi:10.1002/wps.20060
6. Kishimoto T, Robenzadeh A, Leucht C, et al. Long-acting injectable vs oral antipsychotics for relapse prevention in schizophrenia: A meta-analysis of randomized trials. *Schizophr Bull*. 2014;40(1):192-213. doi:10.1093/schbul/sbs150
7. Subotnik KL, Casaus LR, Ventura J, et al. Long-acting injectable risperidone for relapse prevention and control of breakthrough symptoms after a recent first episode of schizophrenia a randomized clinical trial. *JAMA Psychiatry*. 2015;72(8):822-829. doi:10.1001/jamapsychiatry.2015.0270
8. Kane JM, Schooler NR, Marcy P, et al. Effect of Long-Acting Injectable Antipsychotics vs Usual Care on Time to First Hospitalization in Early-Phase Schizophrenia: A Randomized Clinical Trial. *JAMA*

*Psychiatry*. 2020;77(12):1217-1224. doi:10.1001/jamapsychiatry.2020.2076
9. San L, Arranz B, Martinez-Raga J. Antipsychotic drug treatment of schizophrenic patients with substance abuse disorders. *Eur Addict Res*. 2007;13(4):230-243. doi:10.1159/000104886
10. Green AI, Noordsy DL, Brunette MF, O'Keefe C. Substance abuse and schizophrenia: Pharmacotherapeutic intervention. *J Subst Abuse Treat*. 2008;34(1):61-71. doi:10.1016/j.jsat.2007.01.008
11. Green AI. Treatment of schizophrenia and comorbid substance abuse: Pharmacologic approaches. *J Clin Psychiatry*. 2006;67(SUPPL. 7):31-35.
12. Koola MM, Wehring HJ, Kelly DL. The potential role of long-acting injectable antipsychotics in people with schizophrenia and comorbid substance use. *J Dual Diagn*. 2012;8(1):50-61. doi:10.1080/15504263.2012.647345
13. Machielsen MWJ, Veltman DJ, van den Brink W, de Haan L. Comparing the effect of clozapine and risperidone on cue reactivity in male patients with schizophrenia and a cannabis use disorder: A randomized fMRI study. *Schizophr Res*. 2018;194:32-38. doi:10.1016/j.schres.2017.03.030

### SFigure 1. Flowchart of group compositions and project outline.

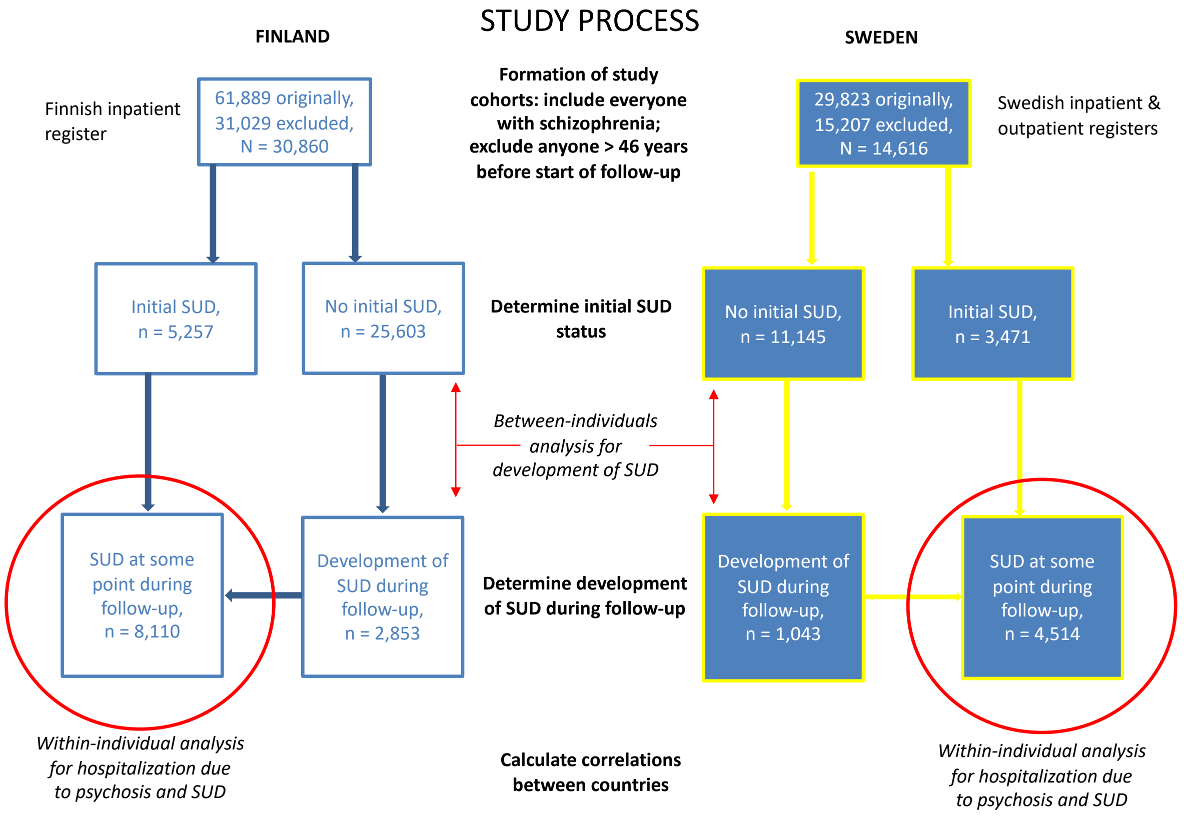

### SFigure 2. Description of within-individual design where each individual act as his/her own control.

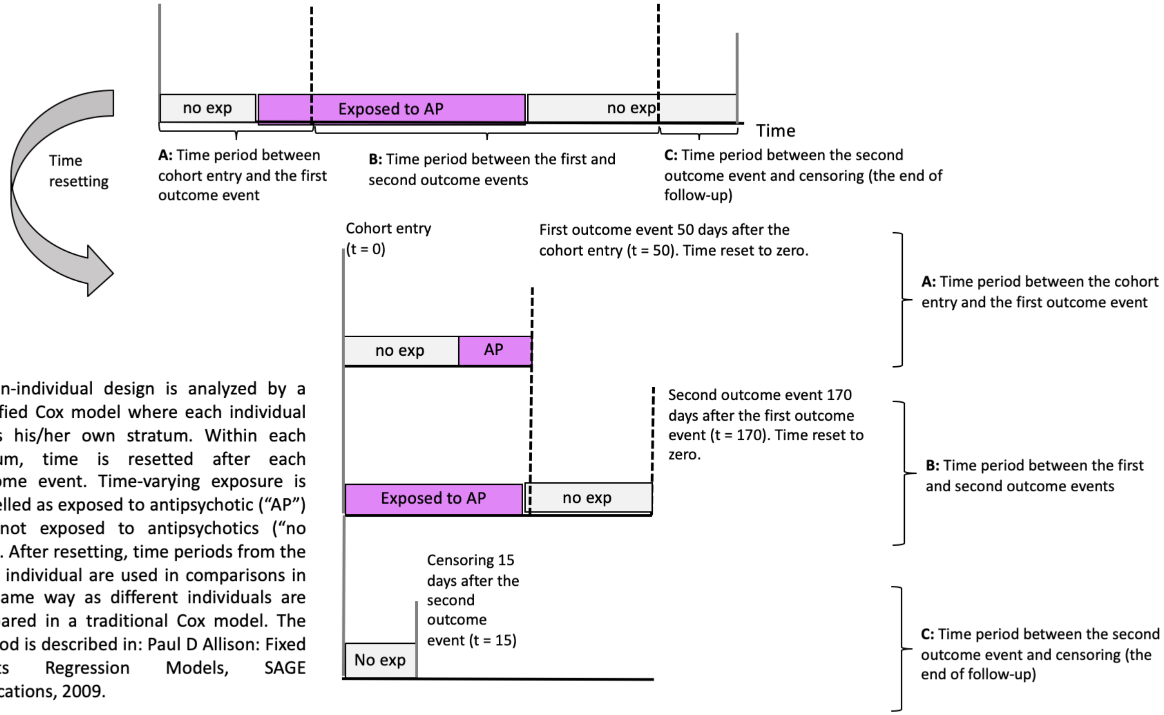


Within-individual design is analyzed by a stratified Cox model where each individual forms his/her own stratum. Within each stratum, time is resetted after each outcome event. Time-varying exposure is modelled as exposed to antipsychotic ("AP") and not exposed to antipsychotics ("no exp"). After resetting, time periods from the same individual are used in comparisons in the same way as different individuals are compared in a traditional Cox model. The method is described in: Paul D Allison: Fixed Effects Regression Models, SAGE Publications, 2009.

**SFigure 3. Risk of developing an initial SUD (among those without SUD) associated with antipsychotic use, between-individuals model in head-to-head comparison to olanzapine users (restricted to cohort ever using olanzapine). A) Finnish cohort N=7755, B) Swedish cohort N=3718. Exposures significant after Benjamini-Hochberg False Discovery Rate correction for multiple comparisons with a 0.05 threshold are bolded. HR= hazard ratio adjusted for covariates.**

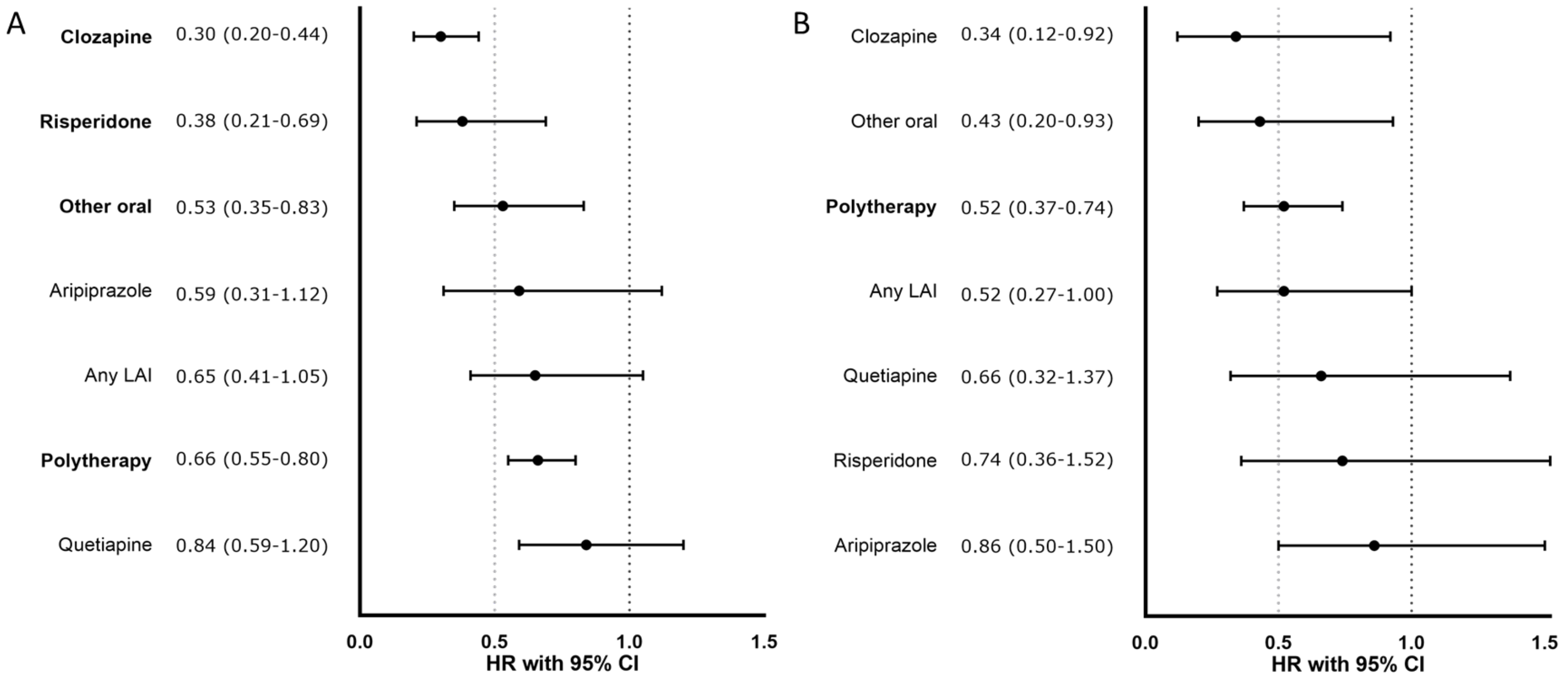

**SFigure 4. Risk of developing an initial SUD (among those without SUD) associated with antipsychotic use with censoring 30 days from beginning of each exposure, between-individuals model (first sensitivity analysis). A) Finnish cohort, B) Swedish cohort. Exposures significant after Benjamini-Hochberg False Discovery Rate correction for multiple comparisons with a 0.05 threshold are bolded. HR= hazard ratio adjusted for covariates.**

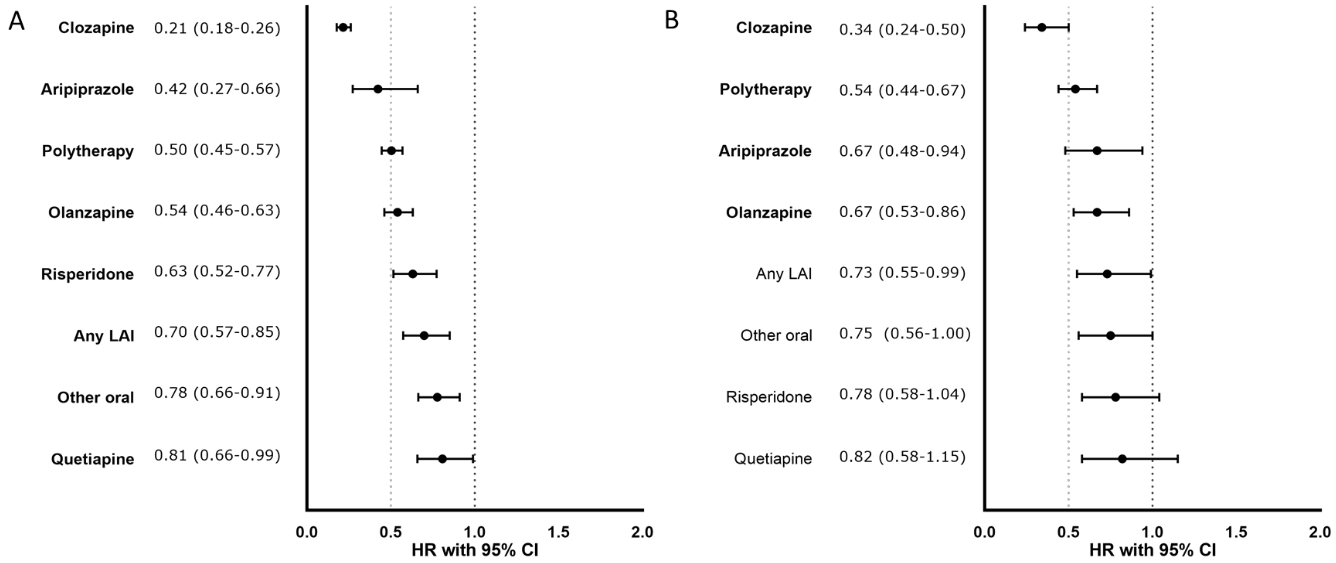

**SFigure 5. Risk of first SUD (among those without SUD) associated with antipsychotic use, excluding patients with a diagnosis of disorganized/hebephrenic schizophrenia, between-individuals model (second sensitivity analysis). A) Finnish cohort, B) Swedish cohort. Exposures significant after Benjamini-Hochberg False Discovery Rate correction for multiple comparisons with a 0.05 threshold are bolded. HR= hazard ratio adjusted for covariates.**

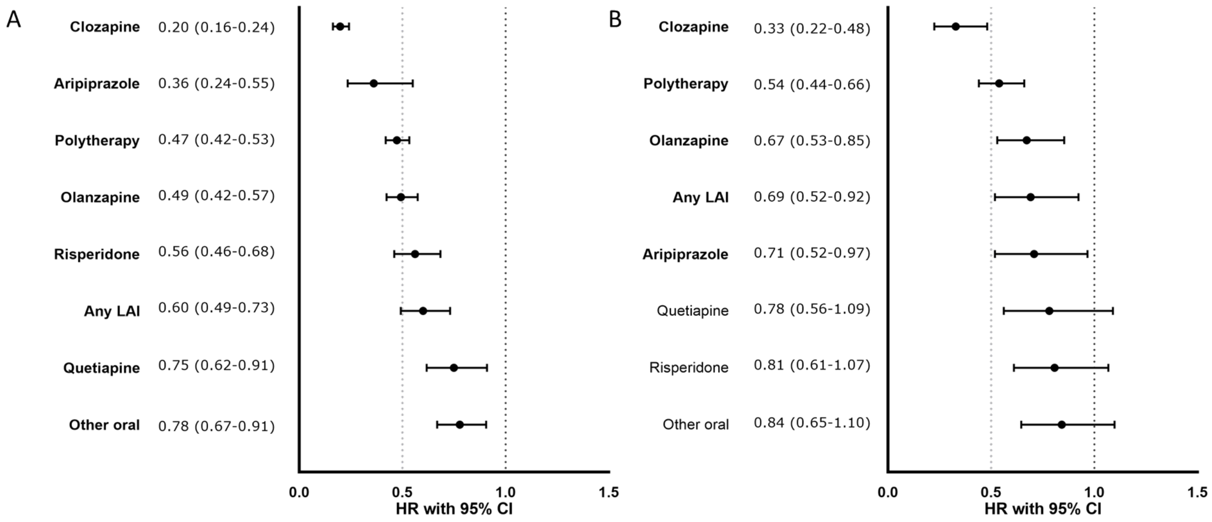

**SFigure 6. Risks of first SUD (among those without SUD) associated with use of specific antipsychotics, between-individuals model stratified by calendar years. A) Finnish cohort, stratified by 1996-2006 and 2007-2017, B) Swedish cohort, stratified by 2006-2011 and 2012-2016. HR= hazard ratio adjusted for covariates. For the Swedish cohort, HR for aripiprazole could not be calculated due to sparsity of data.**

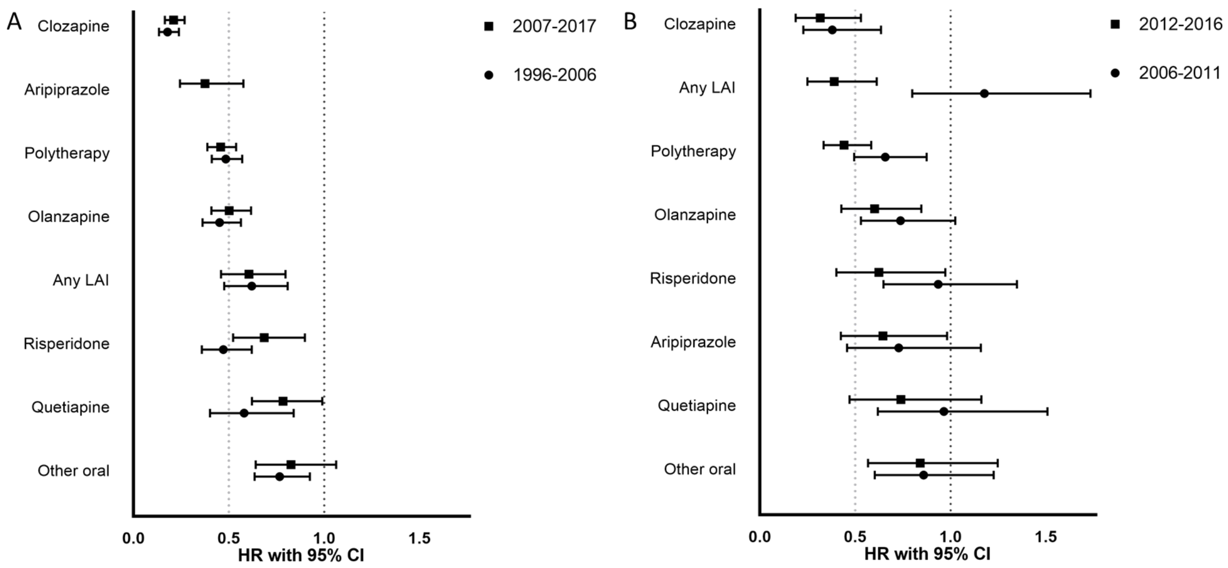

**STable 1. Events and person-years cumulated in the analyses for risk of development of initial SUD in those without SUD, risk of psychiatric hospitalization in those with SUD and risk of SUD hospitalization in those with SUD according to specific antipsychotics**. AP=antipsychotic; SUD=substance use disorder, LAI=long-acting injectable antipsychotic.

| | **Finnish cohort** | | **Swedish cohort** | |
|---|---|---|---|---|
| | Events | Person-years | Events | Person-years |
| **Risk of development of initial SUD in those without SUD** | | | | |
| No antipsychotic | 733 | 76351 | 293 | 17820 |
| Clozapine | 149 | 57704 | 35 | 9323 |
| Olanzapine | 272 | 38302 | 107 | 11719 |
| Quetiapine | 137 | 11271 | 48 | 2817 |
| Risperidone | 132 | 16886 | 67 | 6090 |
| Aripiprazole | 23 | 3538 | 50 | 4842 |
| AP Polytherapy | 826 | 130044 | 243 | 24166 |
| Other oral AP | 272 | 32775 | 80 | 6853 |
| Any LAI | 166 | 20072 | 81 | 7668 |
| **Risk of psychiatric hospitalization in those with SUD** | | | | |
| No antipsychotic | 7512 | 21560 | 4810 | 7386 |
| Clozapine | 3996 | 11832 | 736 | 2310 |
| Olanzapine | 2400 | 7613 | 1028 | 3060 |
| Quetiapine | 1321 | 4196 | 465 | 1116 |
| Risperidone | 981 | 2796 | 450 | 1440 |
| Aripiprazole | 307 | 779 | 500 | 1177 |
| AP Polytherapy | 16367 | 32176 | 6625 | 11213 |
| Other oral AP | 2045 | 5312 | 990 | 1955 |
| Any LAI | 1631 | 4357 | 1597 | 3172 |
| **Risk of hospitalization due to SUD, in those with SUD** | | | | |
| No antipsychotic | 5258 | 21411 | 3858 | 7241 |
| Clozapine | 1064 | 12203 | 166 | 2313 |
| Olanzapine | 1429 | 7711 | 793 | 3054 |
| Quetiapine | 934 | 4222 | 351 | 1119 |
| Risperidone | 613 | 2800 | 260 | 1439 |

| | | | | |
|---|---|---|---|---|
| Aripiprazole | 147 | 780 | 211 | 1180 |
| AP Polytherapy | 6553 | 33204 | 2661 | 11294 |
| Other oral AP | 1345 | 5336 | 637 | 1961 |
| Any LAI | 913 | 4407 | 919 | 3175 |

**SFigure 7. Correlation of effectiveness of antipsychotic treatments (i.e. of adjusted hazard ratios, aHRs) in reducing risk for hospitalization due to SUD in patients with schizophrenia and comorbid SUD between the Finnish and Swedish cohorts, Pearson's r=0.71, p=0.049, N=8 antipsychotics. The linear regression curve is depicted with a red line and the 95% confidence interval area for the curve with dark grey. X-axis: aHRs in Finland. Y-axis: aHRs in Sweden.**

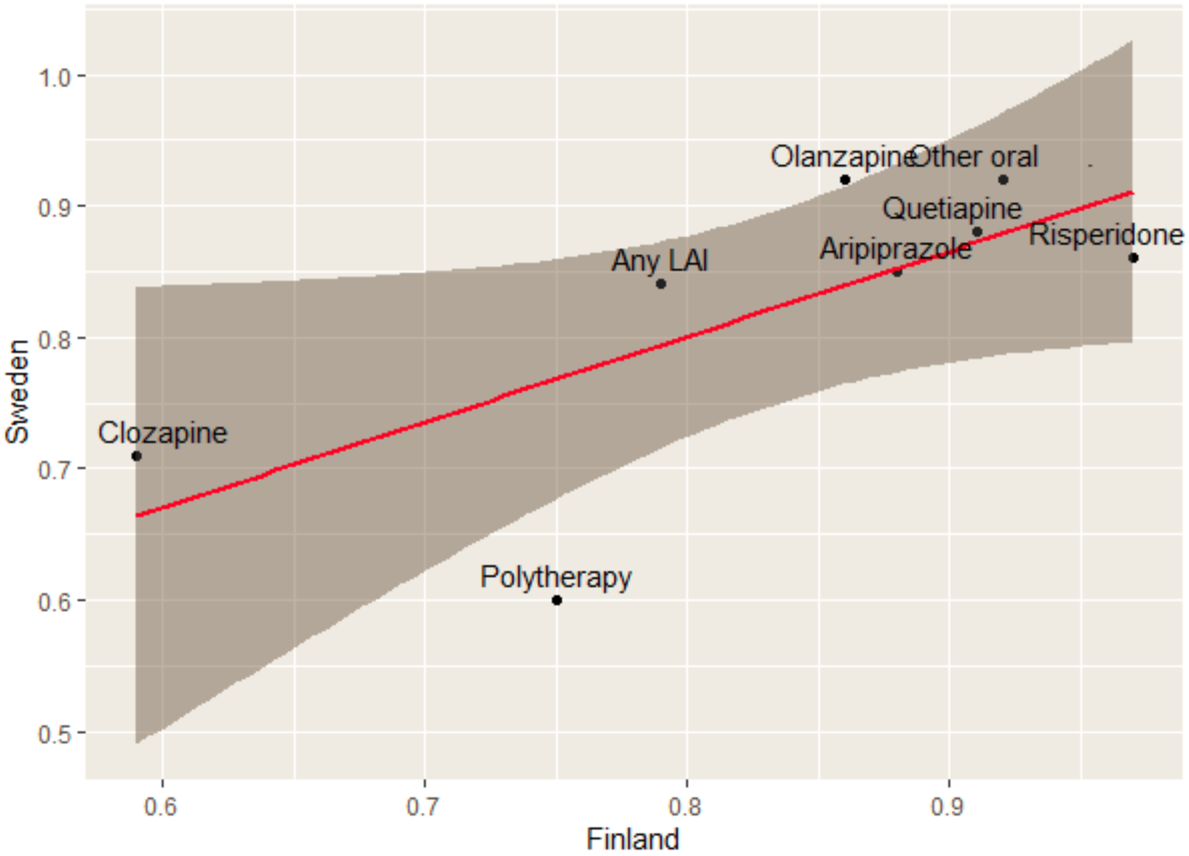
